# What multiple Mendelian randomization approaches reveal about obesity and gout

**DOI:** 10.1101/2021.03.26.21254420

**Authors:** Charleen D. Adams, Brian B. Boutwell

**Affiliations:** Department of Environmental Health, Program in Molecular and Integrative Physiological Sciences, Harvard T.H. Chan School of Public Health, Boston, Massachusetts 02115, USA; School of Applied Science, The University of Mississippi P.O. Box 1848, University, MS, 38677, USA; John D. Bower School of Population Health, University of Mississippi Medical Center, Jackson, MS, 39216, USA

## Abstract

**Background/Objectives:** Gout is a painful arthritic disease. A robust canon of observational literature suggests strong relationships between obesity, high urate levels, and gout. But findings from observational studies can be fraught with confounding and reverse causation. They can conflict with findings from Mendelian randomization (MR), designed to tackle these biases. We aimed to determine whether the relationships between obesity, higher urate levels, and gout were causal using multiple MR approaches, including an investigation of how other closely related traits, LDL, HDL cholesterol, and triglyceride levels fit into the picture.

**Subjects/Methods:** Summary results from genome-wide association studies of the five above-mentioned traits were extracted and used to perform two-sample (univariable, multivariable, and two-step) MR and MR mediation analysis.

**Results:** Obesity increased urate (beta=0.127; 95% CI=0.098, 0.157; *P*-value=1.2E-17) and triglyceride levels (beta=0.082; 95% CI=0.065, 0.099; *P*-value=1.2E-21) and decreased HDL cholesterol levels (beta=-0.083; 95% CI=-0.101, −0.065; *P*-value=2.5E-19). Higher triglyceride levels increased urate levels (beta=0.198; 95% CI=0.146, 0.251; *P*-value=8.9E-14) and higher HDL levels decreased them (beta=-0.109; 95% CI=-0.148, −0.071; *P*-value=2.7E-08). Higher urate levels (OR=1.030; 95% CI=1.028, 1.032; *P*-value=1.1E-130) and obesity caused gout (OR=1.003; 95% CI=1.001, 1.004; *P*-value=1.3E-04). The mediation MR of obesity on gout with urate levels as a mediator revealed, however, that essentially all of the effect of obesity on gout is mediated through urate. The impact of obesity on LDL cholesterol was null (beta=-0.011; 95% CI=-0.030, 0.008; *P*-value=2.6E-01), thus it was not included in the multivariable MR. The multivariable MR of obesity, HDL cholesterol, and triglycerides on urate levels revealed that obesity has an effect on urate levels even when accounting for HDL cholesterol and triglyceride levels.

**Conclusions:** Obesity impacts gout indirectly by influencing urate levels and possibly other traits, such as triglycerides, that increase urate levels. Obesity’s impact on urate is exacerbated by its apparent ability to decrease HDL cholesterol.

## Introduction

Human diets, spanning from ancestral to current, involve substantive consumption of foods which can increase a waste product of human metabolism known as uric acid^1^. This is owed largely to the prevalence of meat and fish (which contain purines), but also to the consumption of fructose and glucose rich foods^1^. The possible downstream consequences of elevated uric acid have interested researchers across fields for some time^1,2^. The most widely examined consequence of uric acid production involves gout, an inflammatory arthritic disease reflecting the accumulation of crystallized urate in joints, most prominently in the largest toe^3^. Correlations exist too, however, between serum uric acid and cardiovascular disease, chronic kidney disease, and metabolic syndrome, an outcome representing a confluence of connected risk factors (e.g., obesity, type 2 diabetes, etc.)^2^. For patients with coronary artery disease, in particular, some associational evidence suggests that uric acid may represent an independent risk factor for mortality^4^. Moreover, a robust canon of epidemiologic literature suggests consistent relationships between obesity, uric acid levels, and gout^5–10^ 5-10. Using the National Health and Nutrition Surveys (NHANES) data on adult participants in the US from 1988-1994 and 2007-2010, for instance, researchers estimated the prevalence of gout among participants of normal BMI to be 1-2%, but 4-5% in those classified with class I obesity and 5-7% in those with class II or III obesity 10. In a population-based study of men in Japan, visceral fat accumulation was identified in 56% of men with hyperuricemia, hinting at the role of obesity in potentially causing higher urate levels^7^.

Complicating research on this topic is the common inability to disentangle cause from effect^3^. Epidemiological designs can provide insight about risk, yet accounting for potential reverse causation, as well as non-random selection across environmental, lifestyle, dietary, and genetic risk factors produce a range of challenges, none easily surmounted^11^. Mendelian randomization (MR), however, which utilizes genome-wide associational (GWA) studies to build genetic “instruments” for putative environmental risk factors—like uric acid—which offer a way to overcome many of these difficulties.

In a recent study of type 2 diabetes (T2D), for example, Sluijs and colleagues (2015) failed to find a causal effect of uric acid levels on T2D when utilizing MR designs, despite replicating the previously apparent epidemiological correlations^3^. The analysis by Sluijs *et al*. (2015) is an important step in better understanding one of the possible health risks imposed by elevated uric acid levels. Still, several interesting hypotheses remain. Elevated uric acid, for example, might be “an effect” of other metabolic disorders—such as obesity—rather than a straightforward causal agent. Here we combine several MR designs, including two-step and multivariable MR to zoom in on the roles of obesity, uric acid, and gout. We conceive of the analysis as having four parts:

1. First, we examine the univariable effects of obesity on four traits: circulating levels of HDL cholesterol, LDL cholesterol, triglycerides, and urate.
2. Second, we examine the univariable effects of three trait-levels (HDL cholesterol, triglycerides, and urate) on gout.
3. Third, we perform a multivariable MR analysis of obesity, HDL cholesterol, and triglycerides in relation to urate levels.
4. Fourth, we perform a MR mediation analysis of the total, direct, and indirect effects of obesity on gout conceiving of urate as the mediator.

## Methods

### Conceptual approach

MR is an instrumental variables technique and analytic approach. It uses insights from Mendel’s laws of inheritance and genotype assignment at conception^12–14^. to improve causal inference in epidemiologic studies. It does this by using summary statistics for genetic variants reliably associated with traits of interest in statistical models instead of the traits themselves (i.e., genetic variants, typically single-nucleotide polymorphisms, SNPs, are used as “instruments”). Doing so prevents much of the confounding and reverse causation that bias estimates in observational studies, so long as certain assumptions are not violated.

### MR assumptions

Key MR assumptions include the following (see ^12–14^):

1. The SNPs serving as genetic instruments must be reliably associated with the exposure of interest.
2. The instrumental SNPs must not be associated with confounders of the exposure-outcome relationship.
3. There must not be any other pathway from the SNP to the outcome other than through the exposure (no pleiotropy).

Before genome-wide association (GWA) studies became common, and prior to their ability to recruit and retain tens, or hundreds, of thousands of participants, MR was performed using SNP data from a single GWA study. It was subsequently adapted to permit extracting SNP data in the form of summary statistics from two separate GWA studies. This adaptation, called two-sample MR is what is used here.

In two-sample MR, summary statistics (e.g., effect estimates, standard errors, and *P*-values) are the data sources^15–20^. Readers less familiar with techniques in genetic epidemiology may confuse individual genotype calls with summary data. To ensure there is not confusion, the effect estimates referred to as “summary statistics” are not the genotypes for individuals but the effects for the associations between the SNPs and the phenotypes in the population of individuals studied. They are the results. Using a two-sample MR design to calculate the causal effect of obesity on gout, for example, estimates of the SNP-obesity associations (*β*^ZX) are calculated in sample 1 (from a GWA study of obesity). The association between these same SNPs and gout is then estimated in sample 2 (*β*^ZY) (from a GWA study of gout). These estimates are combined into Wald ratios (*β*^XY=*β*^ZYβ^ZX). The *β*^XY estimates are meta-analyzed using the inverse-variance weighted (*β*^IVW) method and various sensitivity estimators. The IVW method produces an overall causal estimate for obesity on gout.

In the present study, summary statistics were downloaded from MR-Base (http://www.mrbase.org/ ^15^), a repository of freely available GWA study results (see Supplementary tables 10-12 and 14-18). Descriptions of the GWA studies are provided in the sections below that discuss the MR analyses they were used for.

### Overview of designs

We utilize a variety of two-sample MR designs for the present analysis:

- Univariable MR for testing the effects of:
  - Obesity on urate
  - Obesity on HDL
  - Obesity on LDL
  - Obesity on triglycerides
  - HDL on urate
  - Triglycerides on urate
  - Urate on gout
  - Obesity on gout
- Two-*step* MR (not to be confused with two-*sample* MR, which refers to the data sources; two-step MR) is an approach for testing a hypothesized mechanism. Two-step MR refers to performing two MR analyses sequentially, such that the outcome variable of the first MR analysis is the exposure variable for the second MR analysis. The goal is to take the MR procedure a step beyond knowledge of whether two variables are causally related and to infer a possible mechanism. Here we are interested in knowing whether obesity impacts gout through causing changes to urate levels. The proposed mechanism, therefore, is a pathway from obesity to gout through urate. To explore this, we first instrumented obesity and tested the impact of obesity on urate levels (with a univariable MR analysis of obesity on urate). This is the first “step” in this two-step MR. For the second “step”, we instrumented urate levels and tested the impact of more urate on gout (univariable MR of urate on gout). The reasoning is this: If obesity influences urate and urate influences gout, then urate possibly mediates the relationship between obesity and gout. “Mediation” connotes the presence of a putative mechanism. Two additional two-step MR analyses are likewise performed (see **Figure 1**):

**Fig. 1.**
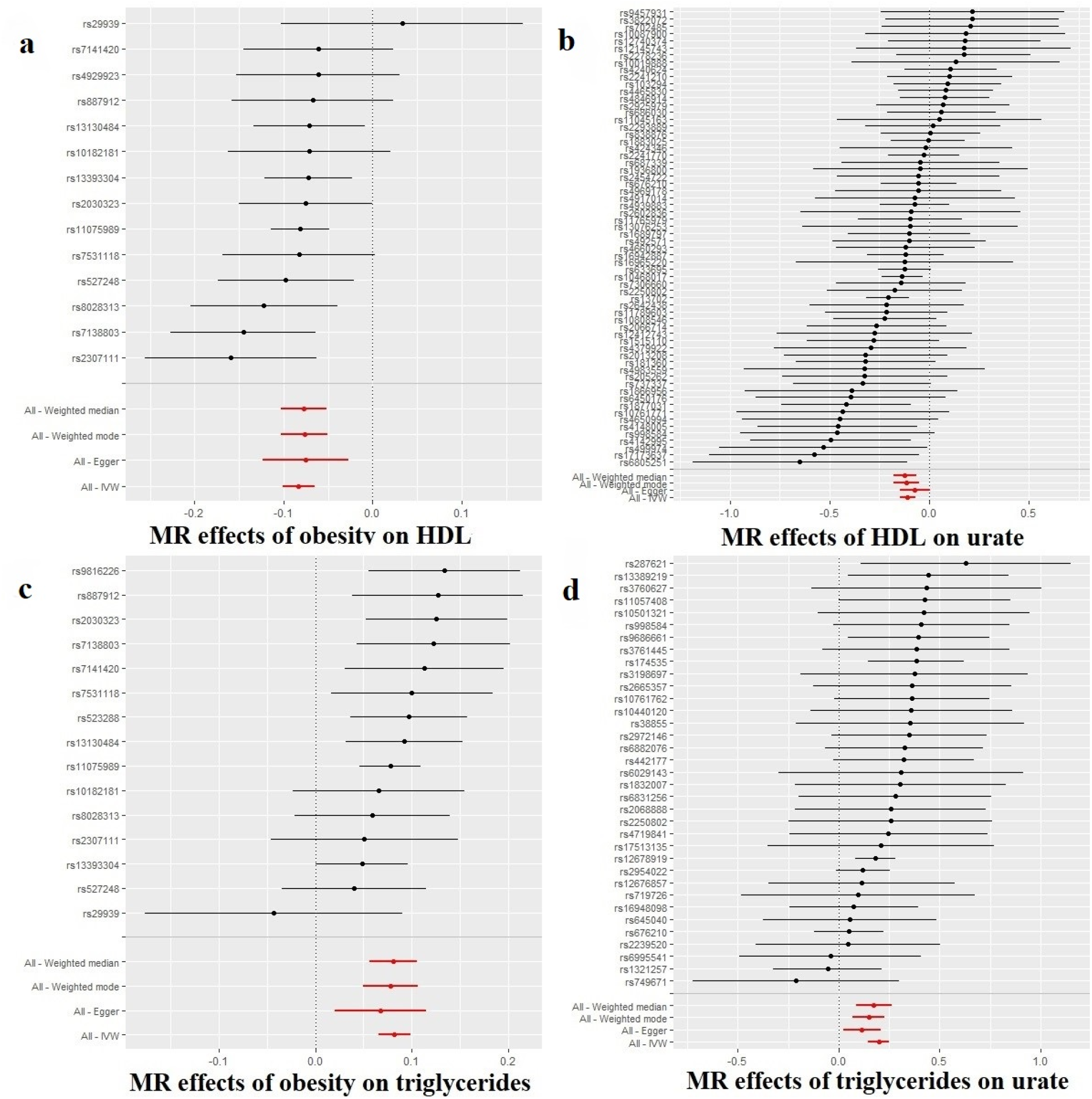
Individual-SNP and meta-analyzed MR results (beta estimates and 95% confidence intervals) for the overall effect of obesity on HDL cholesterol levels (a), the effect of HDL cholesterol levels on urate levels (b), obesity on triglyceride levels (c) and triglyceride levels on urate levels (d). These figures suggest that obesity has impacts on gout, in part, by decreasing HDL cholesterol levels, which reduce urate levels, and by increasing triglyceride levels, which increase urate levels. The effects of obesity on HDL cholesterol levels (a) and HDL cholesterol levels on urate levels (b) comprise a two-step MR analysis treating HDL cholesterol informally as a mediator of obesity on urate levels. The effects of obesity on triglyceride levels (c) and triglyceride levels on urate levels (d) comprise a two separate two-step MR analysis treating triglycerides informally as a mediator of obesity on urate levels.
  - obesity on HDL cholesterol levels and HDL cholesterol levels on urate levels
  - obesity on triglyceride levels and triglyceride levels on urate levels
- Multivariable MR (includes more than one independent variable in the MR model). Multivariable MR permits statistical adjustment, similar to multivariable regression.
- MR mediation analysis (a combination of univariable and multivariable MR approaches) is a complement to two-step MR and used to test a hypothesized mechanism. MR can borrow and incorporate well-established techniques from observational epidemiology. As already eluded to, one of these is mediation analysis. Two-step MR and mediation analysis are complementary approaches for testing pathways/mechanisms. Here we complement the two-step MR of obesity on urate and urate on gout with a mediation analysis to determine the indirect effect of obesity that occurs through obesity’s impact on urate. Three parameters are usually estimated in epidemiologic mediation analyses: i) the total effect (the overall effect of an exposure on an outcome through all potential pathways), ii) the direct effect (the effect of the exposure on an outcome not due to a mediator of interest), and iii) the indirect effect (the effect of exposure on the outcome due to the mediator)^21^. In the context of MR, mediation analysis can be done by generating the total effect with univariate MR (e.g., the effect of obesity on gout). The direct effect can be obtained with multivariate MR (e.g., the effect of obesity on gout when adjusting for urate levels). The indirect effect (i.e., the effect on risk for gout that occurs through the mediator, i.e., increased urate levels) can be calculated by either subtracting the direct effect from the total effect or by multiplying the effect of the exposure on the mediator by the effect of the mediator on the outcome, using the product of coefficients approach^21^. We used the product of coefficients approach. Typically, alongside the total, direct, and indirect effects, a proportion mediated can also be calculated, except when there is evidence for “inconsistent mediation” (a scenario in which the direct and indirect effects are in the opposite directions)^22^. The present MR mediation results (discussed under Results) are an example of inconsistent mediation (i.e., the direct effect of obesity on gout and the indirect effect of obesity on gout are in opposite directions). Thus, the proportion mediated is not calculated.

### Data sources for obesity on urate

#### Instrument data source: obesity

SNPs for a multi-SNP instrument for obesity were extracted from Berndt *et al*. (2013), which performed a meta-analysis of GWA studies of clinically defined obesity in 98 679 participants of European ancestry, of which 32 858 were classified as obese^23^. Obesity case status was defined as BMI ≥ 30 kg/m^2^. Analyses accounted for sex and used a genomic control.

#### Outcome data source: serum urate levels

The outcome data for circulating urate (mg/dl) was extracted from Köttgen *et al*., which performed a meta-analysis of GWA studies for serum urate levels in 110 347 participants of European ancestry^24^. The studies included in the meta-analysis were adjusted for age and sex, as well as study-specific covariates, where applicable (e.g., principal components and study center).

### Data sources for obesity on HDL levels

#### Instrument data source: obesity

The same GWA data source as for obesity ^23^ on urate was used to instrument obesity.

#### Outcome data source: HDL levels

The outcome data for circulating HDL levels (a continuous measure) come from Willer *et al*. (2013), which performed an age- and sex-adjusted GWA study of circulating HDL levels in up to 187 167 individuals, largely of European ancestry. They adjusted for population structure^25^.

### Data sources for obesity on LDL levels

#### Instrument data source: obesity

The same GWA data source as for obesity ^23^ on urate was used to instrument obesity.

#### Outcome data source: LDL levels

The outcome data for LDL levels (a continuous measure) come from Willer *et al*. (2013), which performed an age- and sex-adjusted GWA study of circulating LDL levels in up to 173 082 individuals, largely of European ancestry. They adjusted for population structure ^25^.

### Data sources for obesity on triglyceride levels

#### Instrument data source: obesity

The same GWA data source as for obesity 23 on urate was used to instrument obesity.

#### Outcome data source: triglyceride levels

The outcome data for triglyceride levels (a continuous measure) come from Willer *et al*. (2013), which performed an age- and sex-adjusted GWA study of circulating triglyceride levels in up to 177 861 individuals, largely of European ancestry. They adjusted for population structure ^25^.

### Data sources for HDL on urate levels

#### Instrument data source: HDL levels

The same GWA data source as for HDL levels in the MR of obesity on HDL levels ^25^ was used to instrument HDL.

#### Outcome data source: urate levels

The same GWA data source for urate levels in the MR of obesity on urate levels ^23^ was used for the outcome data.

### Data sources for triglyceride levels on urate levels

#### Instrument data source: triglyceride levels

Same as for the urate levels in the MR of obesity on triglyceride levels ^25^.

#### Outcome data source: urate levels

The same GWA data source for urate levels in the MR of obesity on urate levels ^23^ was used for the outcome data.

### Data sources for urate levels on gout

#### Instrument data source: serum urate levels

The multi-SNP instrument for urate levels (mg/dl) was extracted from Köttgen *et al* (2013)^24^.

#### Outcome data source: self-reported gout

The outcome data for presence of gout was extracted from a GWA study performed by the Medical Research Council-Integrative Epidemiology Unit (MRC-IEU) staff, using PHESANT-derived^26^ UK Biobank data^27,28^ (UK Biobank data field 20002). UK Biobank contains data on ∼500 000 participants, largely of European descent^29^. The MRC-IEU staff adjusted for sex and genotyping chip, and used k-means cluster analysis for European ancestry (first four principal components, as provided by the UK Biobank)^30^. The GWA study contained 462 933 participants, of which 6542 were classified as having self-reported gout.

### Data sources for obesity on gout

#### Instrument data source: obesity

The same GWA data source as for obesity^23^ on urate was used to instrument obesity.

#### Outcome data source: self-reported gout

Same as for the MR of urate levels on gout.

### Instrument construction

For all the approaches, an “instrument” had to be constructed. The summary statistics for the SNPs strongly associated with the exposure traits are the instruments. As mentioned above, the reader can view these in Supplemental tables 10-12 and 14-18.

Independent (those not in linkage disequilibrium, LD; R^2^ < 0.001) SNPs associated at genome-wide significance (*P* < 5 × 10^−8^) with exposure traits were extracted from GWA studies for the exposures in this analysis. The summary statistics for the exposure-associated SNPs were then extracted from each of the outcome GWA studies. SNP-exposure and SNP-outcome associations were harmonized and combined with the inverse variance-weighted (IVW) method using first-order weights (**Fig. 1**).

### Sensitivity analyses

The IVW estimator can be biased in the presence of pleiotropy^31^. To address this, we included a battery of sensitivity estimators, including MR-Egger regression, weighted median, and weighted mode MR methods. The directions and magnitudes of their effect estimates were compared with those of the IVW^32^. If the IVW and sensitivity estimators comport, this provides a qualitative screen against pleiotropy; since the various MR estimators make different assumptions, in the presence of substantial pleiotropy, their estimates are unlikely to align^31,33,34^.

Of note: MR-Egger regression, in addition to providing a (sensitivity) estimate for a causal effect, includes a formal test for directional pleiotropy (the “MR-Egger intercept test”). When the MR-Egger intercept is consistent with 0 (or 1 when exponentiated; *P*>0.05), this provides some evidence against pleiotropy in the IVW estimate. Certain assumptions for MR-Egger regression must hold, however, for this to be true. MR-Egger requires there be negligible measurement error in the SNP-exposure estimates^35^. Violations to this can dilute the MR-Egger estimate and make the MR-Egger intercept test a potential false positive. *I*^2^ statistics are used to check for this. For example, when *I*^2^ statistics are close to 90%, this suggests about 10% possible bias due to measurement error. SIMEX correction can be performed if the bias is much greater than 10%, and is recommended in such cases^36^. We calculated *I*^2^ statistics for the univariate models, which suggested minimal bias due to this problem (SIMEX correction was also run nonetheless; see Supplementary tables 1-3 and 5-9).

Lastly, a final measure was taken to ward against pleiotropy. Heterogeneity in the MR effect estimates for SNPs can indicate pleiotropy. As such, potential outlier SNPs were removed, using RadialMR regression^37^, for all univariable models. Due to this, the various MR tests that have the same exposure (i.e., obesity on urate, obesity on triglycerides, obesity on HDL, etc.) may contain differing numbers of SNPs. All instrumental variables included in this analysis have Cochrane’s *Q*-statistic *P*-values indicating no evidence for heterogeneity between SNPs^38^. Heterogeneity statistics and scatter plots are available in Supplementary tables 1-3 and 5-9).

### Multiple testing

Altogether there were eight univariable MR tests (including the three two-step MRs), one multivariable MR, and one formal MR mediation analysis, making 10 tests. To account for multiple testing, a Bonferroni threshold for stringent evidence was set to *P*<0.05/10=0.005. *P*<0.05 was considered nominally significant, and suggestive.

### Software

The IVW and sensitivity estimations were performed in R version 3.6.2 with the “TwoSampleMR” package^15,39^. The multivariable MR analysis was performed using the “mv_multitiple” function for generating IVW estimates, also within the “TwoSampleMR” package, after clumping for LD, harmonizing^19^ and removing outlier SNPs, which had been removed in the univariate analyses. The standard error for the indirect effect was approximated with the delta method 22.

## Results

We provide a paragraph summarizing the results below it, which include sensitivity analyses for pleiotropy, to make it easier for the reader to navigate:

Obesity increased urate and triglyceride levels and decreased HDL cholesterol levels. Higher triglyceride levels increased urate levels, and higher HDL levels decreased them. Higher urate levels and obesity increase risk for gout. The mediation MR of obesity on gout with urate levels as a mediator revealed, however, that essentially all of the effect of obesity on gout is mediated through urate. The impact of obesity on LDL cholesterol was null, thus it was not included in the multivariable MR. The multivariable MR of obesity, HDL cholesterol, and triglycerides on urate levels revealed that obesity has an effect on urate levels even when accounting for HDL cholesterol and triglyceride levels.

### Univariable MR tests

#### Obesity on HDL

Obesity decreased HDL levels (beta=-0.083; 95% confidence interval (CI)=-0.101, −0.065; *P*-value=2.5E-19). The sensitivity estimators aligned in their directions and magnitudes of effects, providing no evidence for pleiotropy.

#### Obesity on LDL

There was a lack of evidence that obesity impacts LDL levels (beta=-0.011; 95% CI=-0.030, 0.008; *P*-value=2.6E-01). The sensitivity estimators were discrepant across their directions of effect, which is suggestive of pleiotropic distortion. Due to this, LDL was not taken forward in the multivariable analyses.

#### Obesity on triglycerides

Obesity increased triglyceride levels (beta=0.082; 95% CI= 0.065, 0.099; *P*-value=1.2E-21). The sensitivity estimators aligned.

#### HDL on urate levels

There was strong evidence that higher HDL decreased urate levels (beta estimate per unit higher HDL level (mg/dl)=-0.109; 95% CI=-0.148, −0.071; *P*-value=2.7E-08). The sensitivity estimators aligned.

#### Triglycerides on urate levels

There was strong evidence that higher triglycerides increased urate levels (beta estimate per unit higher triglyceride level (mg/dl)=0.198; 95% CI=0.146, 0.251; *P*-value=8.9E-14). The sensitivity estimators aligned. The results for the MR-Egger intercept test reveal potential pleiotropy, however: MR-Egger intercept: 0.004; 95% CI=0.000, 0.008; *P*-value=3.8E-02.

#### Obesity on gout

Obesity causally impacted the risk for gout, driving it higher (odds ratio, OR=1.003; 95% CI=1.001, 1.004; *P*-value=1.3E-04). The sensitivity estimators aligned.

#### Obesity on urate levels

Obesity raised urate levels (beta=0.127; 95% CI=0.098, 0.157; *P*-value=1.2E-17). The sensitivity estimators aligned.

#### Urate levels on gout

Higher urate levels increased the odds of gout (OR=1.030; 95% CI=1.028, 1.032; *P*-value=1.1E-130). The sensitivity estimators aligned.

These results for these last three tests are displayed in Fig. 2.

**Fig. 2.**
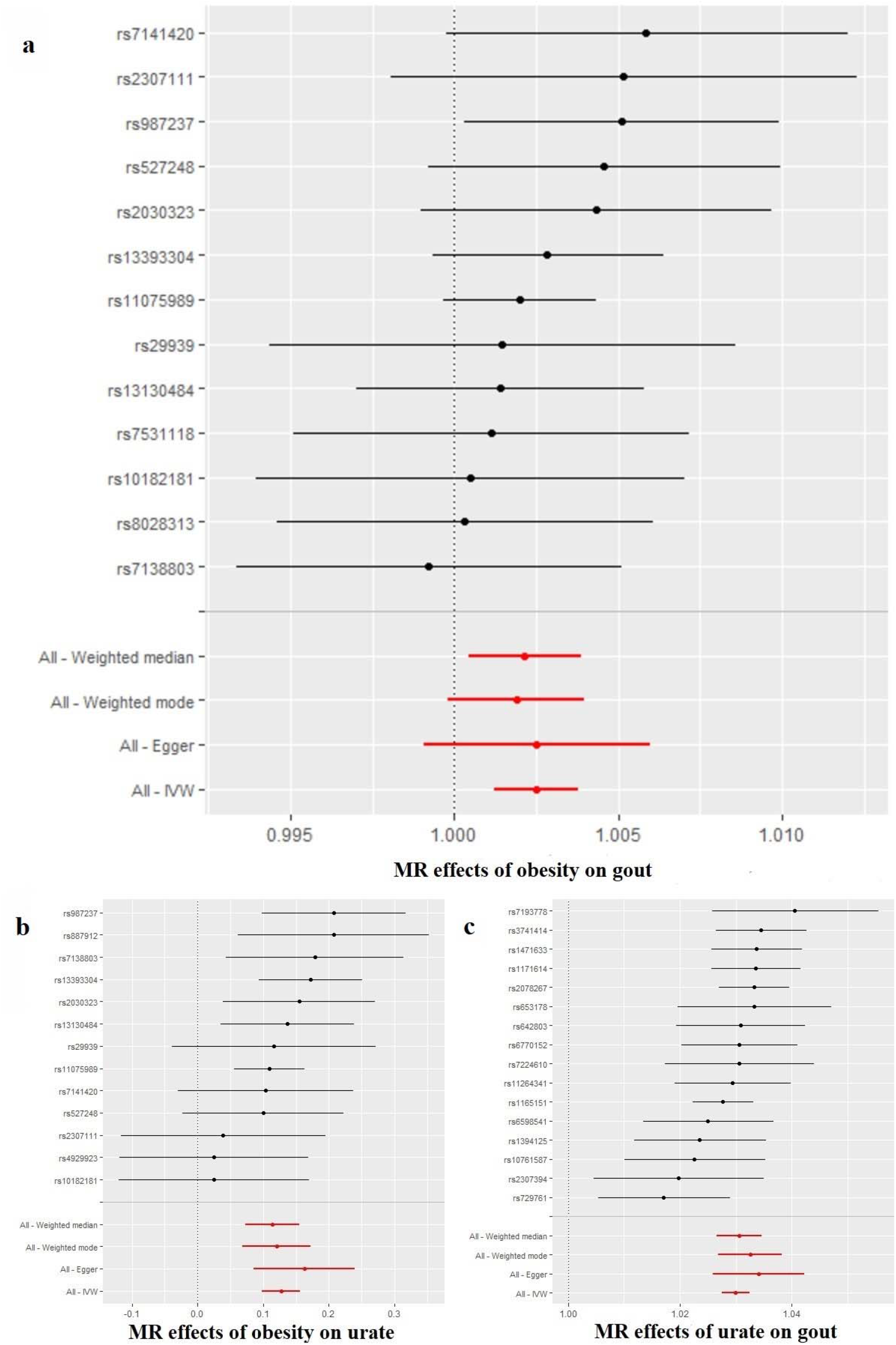
Individual-SNP and meta-analyzed MR results (beta estimates and 95% confidence intervals) for the overall effect of obesity on gout (a), the effect of obesity on urate levels (b) and the effect of urate levels on gout (c). These figures demonstrate that obesity increases urate levels and higher urate levels cause gout. The tests of the effects of obesity on urate levels (b) and urate levels on gout (c) constitute a two-step MR, treating urate informally as a mediator of obesity’s impact on gout.

### Multivariable MR

The multivariable MR of obesity, HDL cholesterol, and triglyceride levels on urate levels revealed strong evidence that obesity and HDL directly impacted urate levels, with obesity leading to higher urate (beta=0.104; 95% CI 0.071, 0.136; *P*-value=4.0E-10) and HDL cholesterol decreasing urate (beta=-0.084; 95% CI −0.130, −0.039; *P*-value=2.8E-04). There was suggestive evidence that higher triglycerides also increased urate levels (beta=0.068; 95% 0.005, 0.132; *P*-value=3.6E-02). See **Table 2**.

**Table 1.**
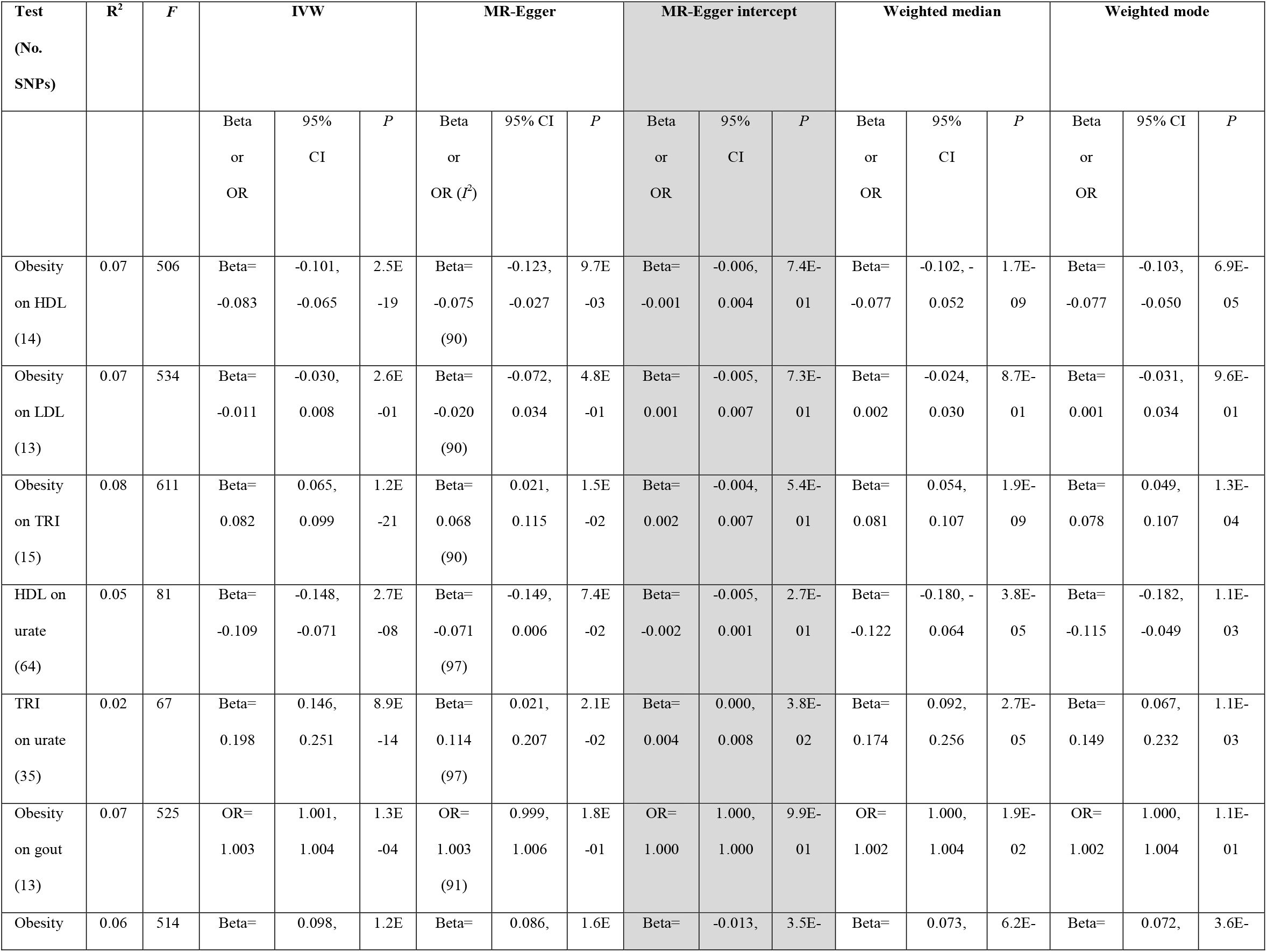
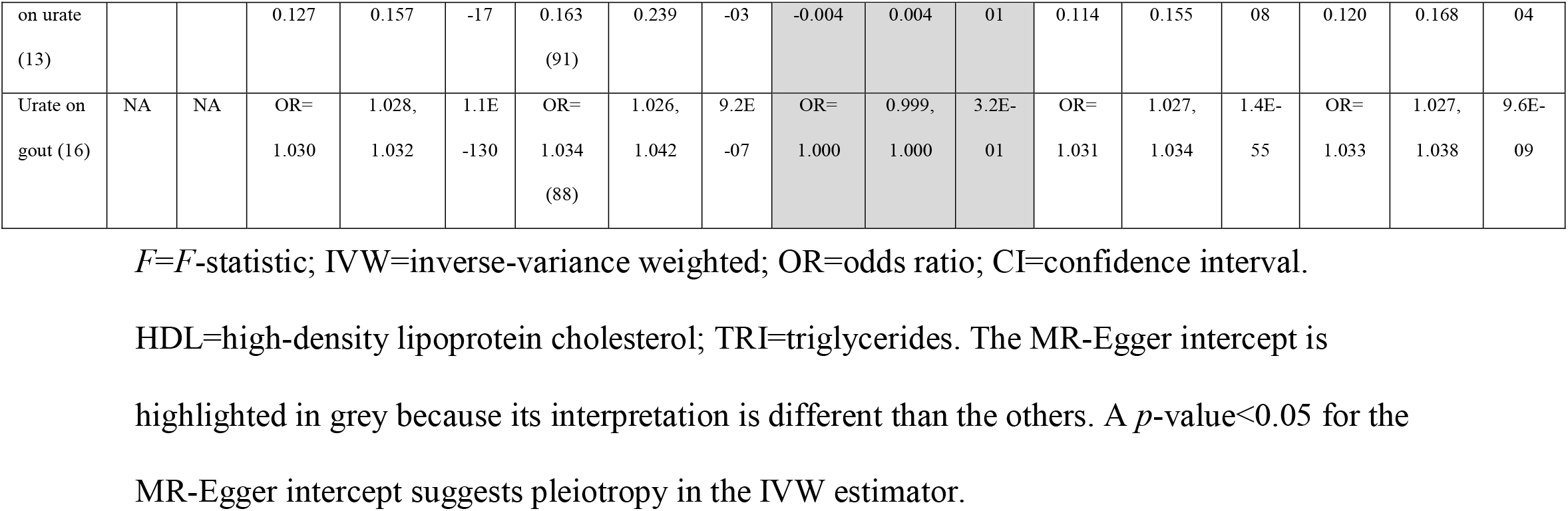
MR results and sensitivity analyses for obesity on gout, obesity on urate levels (Step 1), and urate levels on gout (Step 2).

**Table 2.** Multivariable MR of obesity, HDL cholesterol, and triglycerides on urate levels.

|  | SNP | Beta | Lower 95%<br>CI | Upper 95%<br>CI | P-value |
| --- | --- | --- | --- | --- | --- |
| Obesity | 10 | 0.104 | 0.071 | 0.136 | 4.0E-10 |
| HDL | 62 | -0.084 | -0.130 | -0.039 | 2.8E-04 |
| Triglycerides | 27 | 0.068 | 0.005 | 0.132 | 3.6E-02 |

HDL=HDL cholesterol; CI=95% confidence interval.

### MR Mediation Analysis

The formal mediation analysis of obesity on gout with urate levels as a mediator revealed that nearly all of the effect of obesity on gout is mediated through urate. The reason for this is that the direct effect of obesity on gout is essentially null with the confidence interval including 1 with rounding (OR=0.999; 95% CI 0.997, 1.000; *P*=0.038) whereas the total and indirect effects are not (**Table 3**). (The upper limit for the confidence interval is slightly less than 1 prior to rounding, explaining what looks like, but is not, a discrepancy between the *p*-value and the confidence interval, but the message is the same: the direct of effect of obesity on gout is trivial). Obesity’s impact on gout occurs through factors obesity influences.

**Table 3.**
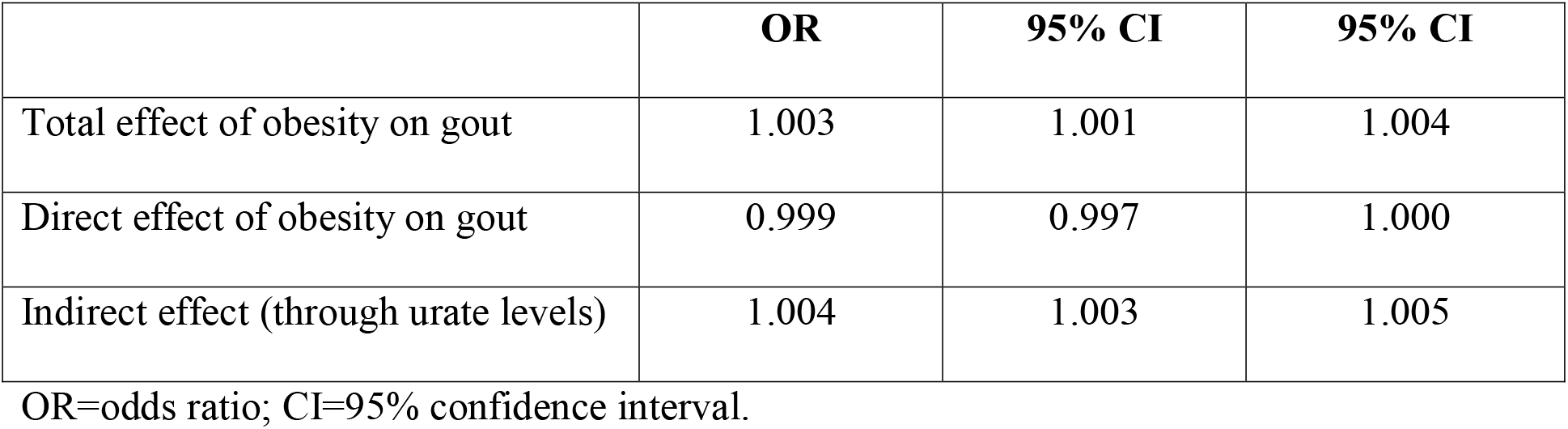
Total, direct, and indirect effects of obesity on gout, with urate levels as the mediator.

|  | OR | 95% CI | 95% CI |
| --- | --- | --- | --- |
| Total effect of obesity on gout | 1.003 | 1.001 | 1.004 |
| Direct effect of obesity on gout | 0.999 | 0.997 | 1.000 |
| Indirect effect (through urate levels) | 1.004 | 1.003 | 1.005 |

OR=odds ratio; CI=95% confidence interval.

## Discussion

Overall, the relationships we investigated suggest that obesity impacts gout through an impact on urate, confirming what has been observed in a robust cannon of epidemiologic literature^8^. MR results can conflict with epidemiologic findings, though, and it was reasonable to think that MR might not replicate the knowledge from the observational literature. Gout exists in a tangled web of metabolic and inflammatory phenotypes, and previous epidemiologic and MR studies have been discrepant with regard to urate’s impact on type 2 diabetes: the epidemiologic data suggest that urate increases risk for T2D, but MR studies (including an analysis we conducted, which is available in Supplementary table 4) imply the epidemiologic association is confounded or reverse caused^3^. In contrast, our findings for the relationship between urate and gout are an example of MR triangulating the previous epidemiologic literature for urate and gout. This helps build a causal story, since causality is not determined from single studies but is a “case” assembled from layers of evidence.

Importantly, from the formal MR mediation analysis, our results suggested that virtually all of the effect of obesity on gout occurred via obesity’s impact on urate (an instance of “inconsistent mediation”). Obesity’s effect on urate, moreover, appears to be occurring by decreasing HDL levels and increasing triglycerides. However, obesity also had a direct effect on urate when accounting for both HDL cholesterol and triglycerides. This suggests that the mechanisms by which obesity impacts urate, and thus gout, are not fully captured by obesity’s impacts on HDL cholesterol and triglycerides. There may be other intermediate traits which obesity influences, which in turn influence urate and gout. Our data provided evidence that this possible third trait is not LDL cholesterol since the impact of obesity on LDL cholesterol did not appear to be causal. Taken as a whole, our results reveal several key points, some previously understood and others less apparent from prior research. While obesity itself is a complex phenotype with various convergent pathways conspiring to produce it, the downstream effects of obesity are also complex and include a variety of deleterious outcomes. Here, we sought to better understand possible pathways leading from obesity to the chronic and painful disorder of gout. When considered as the primary “exposure variable”, obesity appeared to operate antagonistically against protective factors (such as HDL levels), while exacerbating risk factors for gout (urate levels).

Major strengths of our analyses were the use of multiple large GWA studies, capitalizing on their large sample sizes, and the various sensitivity tests we performed to investigate pleiotropy. The only tests that showed evidence for possible pleiotropy were those for the impact of obesity on LDL cholesterol and triglycerides on urate. We did not see evidence for pleiotropy for the relationships between obesity, urate, and gout.

Our primary contribution was to reveal an apparent mediation effect whereby obesity raised uric acid levels, thereby further increasing the likelihood of gout in a causal manner. Indeed, nearly all of the effect of obesity on gout appeared to result from the rise in uric acid. Considered via a translational lens, our results suggest that management of uric acid levels, should help to blunt the effects of obesity on the development of gout.

## Data Availability

All data are freely available through MR-Base.

## Data Availability

All data is publicly available.

https://unstats.un.org/unsd/demographic-social/products/vitstats/seratab2.pdf

https://www.who.int/publications/m/item/weekly-epidemiological-update---19-january-2021

## References

1. Choi, H. K., Liu, S. & Curhan, G. Intake of purine-rich foods, protein, and dairy products and relationship to serum levels of uric acid: the third national health and nutrition examination survey. Arthritis Rheum 52, 283–289 (2005).

2. Sui, X., Church, T. S., Meriwether, R. A., Lobelo, F. & Blair, S. N. Uric acid and the development of metabolic syndrome in women and men. Metabolism 57, 845–852 (2008).

3. Sluijs, I. et al. A Mendelian randomization study of circulating uric acid and type 2 diabetes. Diabetes 64, 3028–3036 (2015).

4. Madsen, T. E. et al. Serum uric acid independently predicts mortality in patients with significant, angiographically defined coronary disease. Am J Nephrol 25, 45–49 (2005).

5. Lee, J. et al. Visceral fat obesity is highly associated with primary gout in a metabolically obese but normal weighted population: A case control study. Arthritis Res Ther 17, 17–79 (2015).

6. Lyu, L.-C. et al. A case-control study of the association of diet and obesity with gout in Taiwan 1-3. Am J Clin Nutr 78, 690–701 (2003).

7. Tamba, S. et al. Relationship between the serum uric acid level, visceral fat accumulation and serum adiponectin concentration in Japanese men. Intern Med 47, 1175–1180 (2008).

8. Thottam, G. E., Krasnokutsky, S. & Pillinger, M. H. Gout and metabolic syndrome: a tangled qeb. Curr Rheumatol Rep 19, 19–60 (2017).

9. Ali, N. et al. Prevalence of hyperuricemia and the relationship between serum uric acid and obesity: A study on Bangladeshi adults. PLoS ONE 13, (2018).

10. Juraschek, S. P., Miller, E. R. & Gelber, A. C. Body mass index, obesity, and prevalent gout in the United States in 1988-1994 and 2007-2010. Arthritis Care Res (Hoboken) 65, 127–132 (2013).

11. Rohrer, J. M. Thinking clearly about correlations and causation: graphical causal models for observational data. Adv Methods Pr Psychol Sci 1, 27–42 (2018).

12. Davey Smith, G. D. & Ebrahim, S. “Mendelian randomization”: can genetic epidemiology contribute to understanding environmental determinants of disease? Int J Epidemiol vol. 32 (2003).

13. Schooling, C. M., Freeman, G. & Cowling, B. J. Mendelian randomization and estimation of treatment efficacy for chronic diseases. Am J Epidemiol 177, 1128–1133 (2013).

14. Hemani, G., Bowden, J. & Davey Smith, G. Evaluating the potential role of pleiotropy in Mendelian randomization studies. Hum Mol Genet 27, R195–R208 (2018).

15. Hemani, G. et al. The MR-Base platform supports systematic causal inference across the human phenome. eLife (2018) doi:10.7554/eLife.34408.

16. Burgess, S., Butterworth, A. & Thompson, S. G. Mendelian randomization analysis with multiple genetic variants using summarized data. Genet Epidemiol 37, 658–665 (2013).

17. Bowden, J., Smith, G. D. & Burgess, S. Mendelian randomization with invalid instruments: effect estimation and bias detection through Egger regression. Int J Epidemiol 44, 512–525 (2015).

18. Johnson, T. Efficient calculation for multi-SNP genetic risk scores. in American Society of Human Genetics Annual Meeting (2012).

19. Davey Smith, G. & Hemani, G. Mendelian randomization: genetic anchors for causal inference in epidemiological studies. Hum Mol Genet 23, (2014).

20. Burgess, S. & Thompson, S. G. Interpreting findings from Mendelian randomization using the MR-Egger method. Eur J Epidemiol 32, (2017).

21. Carter, A. R. et al. Mendelian randomisation for mediation analysis: Current methods and challenges for implementation. bioRxiv (2019) doi:10.1101/835819.

22. MacKinnon, D. P., Fairchild, A. J. & Fritz, M. S. Mediation analysis. Annu Rev Psychol 58, 593–614 (2007).

23. Berndt, S. I. et al. Genome-wide meta-analysis identifies 11 new loci for anthropometric traits and provides insights into genetic architecture. Nat Genet 45, 501–512 (2013).

24. Köttgen, A. et al. Genome-wide association analyses identify 18 new loci associated with serum urate concentrations. Nat Genet 45, 145–154 (2013).

25. Willer, C. J. et al. Discovery and refinement of loci associated with lipid levels. Nat Genet 45, 1274–1285 (2013).

26. Millard, L. A. C., Davies, N. M., Gaunt, T. R., Smith, G. D. & Tilling, K. Software application profile: PHESANT: A tool for performing automated phenome scans in UK Biobank. Int J Epidemio 47, 29–35 (2018).

27. Collins, R. What makes UK Biobank special? Lancet 31, 1173–1174 (2012).

28. Sudlow, C. et al. UK Biobank: an open access resource for identifying the causes of a wide range of complex diseases of middle and old age. PLoS Medicine 12, (2015).

29. Fry, A. et al. Comparison of sociodemographic and health-related characteristics of UK Biobank participants with those of the general population. Am J Epidemiol 186, 1026–1034 (2017).

30. Mitchell, R. H. G. D. T. C. L. H. S. P. L. UK Biobank genetic data: MRC-IEU quality control, version 2. (2019).

31. Spiller, W., Davies, N. M. & Palmer, T. M. Software application profile: Mrrobust - A tool for performing two-sample summary Mendelian randomization analyses. Int J Epidemiol 48, 664–690 (2019).

32. Lawlor, D. A., Tilling, K. & Smith, G. D. Triangulation in aetiological epidemiology. Int J Epidemiol 45, 1866–1886 (2016).

33. Yarmolinsky, J. et al. Appraising the role of previously reported risk factors in epithelial ovarian cancer risk: A Mendelian randomization analysis. PLoS Med 16, (2019).

34. Hwang, L. D., Lawlor, D. A., Freathy, R. M., Evans, D. M. & Warrington, N. M. Using a two-sample Mendelian randomization design to investigate a possible causal effect of maternal lipid concentrations on offspring birth weight. Int J Epidemiol 48, 1457–1467 (2019).

35. Bowden, J. et al. Assessing the suitability of summary data for two-sample mendelian randomization analyses using MR-Egger regression: The role of the I 2 statistic. Int J Epidemiol 45, 1961–1974 (2016).

36. Spiller, W., Slichter, D., Bowden, J. & Davey Smith, G. Detecting and correcting for bias in Mendelian randomization analyses using Gene-by-Environment interactions. Int J Epidemiol 48, 702–707 (2019).

37. Bowden, J. et al. Improving the visualization, interpretation and analysis of two-sample summary data Mendelian randomization via the Radial plot and Radial regression. Int J Epidemiol 47, (2018).

38. del Greco, F., Minelli, C., Sheehan, N. A. & Thompson, J. R. Detecting pleiotropy in Mendelian randomisation studies with summary data and a continuous outcome. Stat Med 34, 2926–2940 (2015).

39. R Core Team. R: a language and environment for statistical computing. (2020).

